# Combined Flywheel Resistance and Aerobic Exercise on Power output and Function in Chronic Kidney Disease: An Exploratory Study on the Influence of Physical Activity

**DOI:** 10.64898/2026.04.14.26350873

**Authors:** Jared M. Gollie, Alice S. Ryan, Michael O. Harris-Love, Peter Kokkinos, Joel D. Scholten, R. Jamil Pugh, Cooper G. Hazel, Marc R. Blackman

## Abstract

**New Findings:** What is the central question of this study? Are adaptations elicited by combined flywheel resistance and aerobic exercise influenced by habitual physical activity levels in patients with chronic kidney disease?

What is the main finding and its importance? Combined flywheel resistance and aerobic exercise promote clinically meaningful improvements in muscle size, power output, and physical function in patients with chronic kidney disease. Those not meeting the weekly moderate intensity physical activity recommendations experienced greater increases in power output compared to those who were physically active whereas no differences in the magnitude of improvements in physical function were observed between physical activity levels.

Physical inactivity is common in chronic kidney disease (CKD) and is associated with poor neuromuscular and functional outcomes. Whether habitual physical activity (PA) influences adaptations to structured exercise in CKD remains unclear. This study examined if adaptations to combined flywheel resistance and aerobic exercise (FRE+AE) differed based on self-reported PA in Veterans with CKD stages 3–4. Twenty older male Veterans with CKD stages 3-4 (mean eGFR 37.9 ± 10.2 mL/min/1.73 m²) were randomized to six weeks of FRE+AE (n=11) or health education (EDU; n=9). Participants were classified as meeting (Meets PA) or below (Low PA) weekly moderate intensity PA recommendations using the 7-day Physical Activity Recall. Outcomes included vastus lateralis muscle thickness (VL MT), knee extensor power output (60°·s^-1^ and 180°·s^-1^), gait speed (GS), and five-repetition sit-to-stand (STS). FRE+AE increased VL MT (p=0.030), power output at 180°/s (p=0.021), GS (p=0.001), and reduced STS time (p=0.012), with significant between-group differences versus EDU for VL MT (p=0.009) and GS (p=0.028). Low PA experienced greater increases in power output at 60°·s^-1^ (Hedge’s g; Low PA=0.44, Meets PA=0.25) and 180°·s^-1^ (Hedge’s g; Low PA=1.38, Meets PA=0.38) compared to Meets PA after FRE+AE. Conversely, Meets PA had greater improvements in GS (Hedge’s g; Low PA=0.93, Meets PA=1.29) and STS (Hedge’s g; Low PA=-0.72, Meets PA=-2.20) compared to Low PA. Six weeks of FRE+AE produced clinically meaningful neuromuscular and functional improvements in Veterans with CKD stages 3-4 irrespective of PA level, supporting FRE+AE as a feasible intervention in this population.

## INTRODUCTION

Physical activity (PA) refers to any bodily movement produced by skeletal muscles that results in an increase in caloric requirements over resting energy expenditure (American College of Sports Medicine *et al*., 2026). PA guidelines for adults recommend engaging in at least 150-300 minutes of moderate intensity aerobic activity or 75-150 minutes of vigorous intensity aerobic activity per week (U.S. Department of Health and Human Services, 2018; American College of Sports Medicine *et al*., 2026). Physical inactivity is associated with an increased risk of chronic, non-communicable diseases (Booth *et al*., 2012). The economic burden of physical inactivity on healthcare systems is substantial, with an estimated $192 billion per year incurred in the United States alone from 2012-2019 (Matjasko *et al*., 2025).

PA status is often considered a confounding variable in exercise studies. The concept of diminishing returns implies that for any given increase in the amount of PA, the magnitude of benefit is inversely associated with baseline activity level (Haskell, 1994). However, as it pertains to chronic aerobic exercise and metabolic adaptations, those with higher habitual PA levels experience the greatest metabolic adaptations (Swift *et al*., 2012; Burton *et al*., 2021; Coyle *et al*., 2022). Whether these findings hold true for neuromuscular and physical function outcomes in response to resistance exercise or to the combination of resistance and aerobic exercise is less clear.

Chronic kidney disease (CKD) is among the top ten leading causes of death and the twelfth leading cause of disability-adjusted life-years worldwide (Mark *et al*., 2025). Adults with CKD are reported to be more sedentary than their non-CKD counterparts (Johansen *et al*., 2000; Beddhu *et al*., 2009). Importantly, low levels of PA in the CKD population are associated with an increased risk for all-cause mortality and cardiovascular disease events (Beddhu *et al*., 2009, 2015; Zelle *et al*., 2011; Rosas *et al*., 2012; Kang *et al*., 2020). Patients with CKD are also at an increased risk for skeletal muscle wasting, neurological impairments, and compromised physical function (Rampersad *et al*., 2021; Wang *et al*., 2022; Arnold *et al*., 2022). The combination of CKD and physical inactivity further exacerbates declines in neuromuscular health and functional capabilities. Although promoting greater uptake of PA in the CKD population has been emphasized (Zelle *et al*., 2017), PA alone may not be sufficient to increase neuromuscular capacity. Therefore, exercise interventions designed to specifically enhance neuromuscular properties in the CKD population are of need.

Flywheel resistance exercise (FRE) uses a cord/strap connected to a rotating shaft that holds a flywheel disc (Tesch *et al*., 2017; Beato *et al*., 2024). As the concentric phase of the exercise movement is initiated, the cord/strap begins to unravel from the shaft, and the flywheel disc(s) begins to rotate. Once the concentric phase of the contraction is complete, the cord/strap rewinds around the fixed shaft requiring the individual to resist the pull of the rotating flywheel disc(s) during the eccentric phase of the stretch-shortening cycle. The unique loading characteristics of flywheel technology allow eccentric output (force and power) to be manipulated by inertial load and contraction speed. FRE is an effective strategy for improving muscle size, strength, and function in younger and older healthy adults (Maroto-Izquierdo *et al*., 2017; Sañudo *et al*., 2019; Hill *et al*., 2022). Moreover, when combined with aerobic exercise, FRE has been shown to elicit greater increases in muscle size and oxidative enzymatic activity without compromising gains in strength when compared to resistance exercise alone (Lundberg *et al*., 2013). These findings make FRE an appealing exercise approach for patients with CKD who are at risk for cardiovascular and neuromuscular complications.

U.S. Veterans constitute a high-risk population for CKD due to their elevated prevalence of the disease and high burden of comorbidities such as diabetes, hypertension, and cardiovascular disease relative to the general population. Current PA and exercise recommendations for adults with CKD align with guidelines for older adults (U.S. Department of Health and Human Services, 2018; Stevens *et al*., 2024; American College of Sports Medicine *et al*., 2026). However, the PA and exercise guidelines for older adults may not be appropriate or attainable for patients with CKD given the sedentary nature of the population and the complexity of the pathophysiology. Furthermore, aerobic exercise has historically been prioritized over resistance exercise in patients with CKD. Whether the benefits of FRE when combined with aerobic exercise (FRE+AE) translate to the CKD population, and if habitual PA levels influence the magnitude of adaptations, have yet to be examined. The purpose of this study, therefore, was to examine the effects of FRE+AE in patients with CKD stages 3-4 and explore potential differences in the magnitude of adaptations based on self-reported PA level. We hypothesized that FRE+AE would promote increases in muscle size, strength, and physical function, and that the magnitude of these adaptations would be greater in patients with lower self-reported PA levels compared to those with higher self-reported PA levels.

## METHODS

### Study Design and Protocol

This study used a prospective, randomized controlled trial comparing six weeks of FRE+AE to health education (EDU) in older male U.S. Veterans with CKD stages 3 & 4. Community-dwelling U.S. Veterans were screened for potential enrollment at the Veterans Affairs Medical Center in Washington D.C. (DC VAMC) from October 2022-September 2025. The study was approved by the DC VAMC Institutional Review Board (IRB) and Research and Development (R&D) Committee prior to the commencement of research activities. Inclusion criteria for study enrollment required participants to be ambulatory with or without use of assistive devices (i.e., cane, walker), aged 50-84 years, diagnosed with CKD stage 3 or 4 defined as estimated glomerular filtration rate (eGFR) 59-15 ml/min/1.73 m^2^ according to Kidney Disease Improving Global Outcomes (KDIGO) guidelines, and not requiring kidney replacement therapy (Stevens *et al*., 2024). Exclusion criteria were a history of acute kidney injury, inability to follow study instructions, peripheral artery disease, and any uncontrolled cardiovascular or musculoskeletal problems that would make study participation unsafe. All study participants voluntarily provided written informed consent using a DC VAMC IRB and R&D approved form before study participation. The data presented herein are part of a larger randomized controlled trial (ClinicalTrials.gov NCT04397159).

Each participant reported to the Skeletal Muscle Laboratory at the DC VAMC to complete neuromuscular strength and physical function assessments lasting 60-90 minutes. Participants were asked to refrain from any vigorous activity for at least 24 hours prior to testing. Testing sequence consisted of diagnostic ultrasound evaluations followed by functional assessments and strength testing.

### 7-Day Physical Activity Recall

The 7-day Physical Activity Recall (PAR) questionnaire was used to assess participants’ PA levels (Hayden-Wade *et al*., 2003). Participants were asked to report activities of the same intensity or greater than walking, performed for at least 10 consecutive minutes. Participants also reported the time spent sleeping each night. Activity intensity was categorized as moderate (similar to walking at a normal pace), very hard (similar to running), or hard (between walking and running). Recall began with the preceding day and then worked backward chronologically until the previous seven days had been accounted for to the best of their ability. The duration and intensity of each bout were recorded, and the totals were summed for the week. Participants were then categorized into those meeting the PA recommendations (≥150 min/week of moderate-intensity activity) and those below the PA recommendations (<150 min/week of moderate-intensity activity).

### Customary Gait Speed (GS)

Customary GS was determined by measuring the time it took each participant to walk 3 meters. Participants were allowed to use ambulatory aids, such as a cane or walker, if desired. Before the test, participants were instructed to walk at their normal walking speed and continue for 2 steps beyond the finish line to prevent deceleration. Participants began each trial with their toes aligned with the starting line and began walking upon receiving the “go” command, at which point the timer was started and continued until both feet crossed the finish line. Two trials were performed, and the faster of the two trials was used for analysis.

### Five Repetition Sit-to-Stand (STS) Test

Participants were asked to perform five repetitions of the STS task as quickly and safely as possible. Participants began by sitting quietly on a seat (height 43 cm) with their back against the backrest of the chair and their hands folded across their chest. If the participant was able to complete one STS repetition successfully without using his arms, he was provided the following standardized instructions for completing the test: “Please stand up straight as quickly as you can five times, without stopping in between. After standing up each time, sit down and touch your back to the backrest of the chair, then stand up again, keeping your arms folded across your chest.” Performance in the STS was determined by the time taken to successfully complete all five repetitions.

### Timed-Up and Go (TUG)

Participants began in a seated position with their back against the chair’s backrest and their arms resting on the armrests. Participants were instructed to stand up, walk to a line 3 meters away, turn around, walk back to the chair, and sit down. The test was scored based on the time taken to complete the task, measured with a handheld stopwatch. Time began when the test administrator said “go” and ended when the participants returned and touched the backrest of the chair.

### Strength Assessments

Maximal voluntary isometric torque (MVIT) was determined by asking participants to kick their foot out as hard and as fast as possible and to hold the contraction for 5 seconds for each isometric contraction. Three repetitions were completed and interspersed with 60-second rest intervals in between contractions. Unilateral peak isokinetic knee extension power output (60°·s^-1^ and 180°·s^-1^) was obtained across three sets of five continuous repetitions using a dynamometer (Biodex System 4 Dynamometer, Biodex Medical, Shirley, NY, USA) in a seated position per manufacturer guidelines. The highest peak power output achieved was used for analysis. All formal strength testing was preceded by a familiarization session and submaximal warm-up repetitions. Visual feedback of velocity and torque values were displayed on a computer monitor throughout the duration of the test to provide knowledge of performance and motivation to the participants.

### Quadriceps Muscle Thickness (MT)

Estimates of quadriceps muscle thickness were determined as the sum of the RF and VL of the dominant limb using B-mode ultrasound with a 5-18 MHz linear array transducer (Nobulus, Hitachi Aloka Medical, Parsippany, NJ). Resting ultrasound images were obtained at 33% and 50% of femur length for VL and RF, respectively. Participants were in the supine position with the knee fully extended. Care was taken to ensure as little pressure as possible was applied during imaging. Ultrasound images were measured three times within the fascial borders of the muscle of interest. The region of interest was defined as the area within the superior and inferior fascial borders and the lateral borders. In instances when a portion of a fascial border was poorly visualized, the examiner used the trajectory of the visible fascial border to complete the region of interest selection. All images will be processed offline using ImageJ (version 1.5; National Institutes of Health, Bethesda, MD, USA).

### Body Composition

Body composition measures were obtained via bioelectrical impedance analysis (BIA) (Tanita MC-980U plus). Analyzed measures included body mass, body mass index (BMI), fat mass (FM), and fat-free mass (FFM). Participants were instructed to avoid strenuous activity for 24 hours prior to testing and asked to limit their water intake. BIA assessment was performed before any physical function or strength assessments to avoid the impact that such activities may have on BIA measures.

### Exercise Intervention

Participants assigned to the FRE+AE group reported to DC VAMC twice a week for six weeks to complete their exercise sessions under the direct supervision of a trained exercise physiologist. Each exercise session consisted of a five-minute warm-up followed by FRE+AE. FRE included three sets of twelve repetitions for the squat and seated shoulder press exercises (kBox 4, Exxentric, AB, Bromma, Sweden). Each set was separated by 30-60 seconds rest, and each exercise separated by 3-5 minutes rest. Each set consisted of two submaximal repetitions, during which participants worked up to their target movement velocity, followed by 12 maximal repetitions. Customized software (LabView) was built to provide live visual feedback of the flywheel’s rotational velocity on a computer monitor.

FRE consisted of a familiarization period (weeks one and two) in which the participants were introduced to the equipment and practiced the squat exercise using the lowest inertial load (0.010 kgm^2^). Participants were then subjected to a range of inertial loads according to their capabilities. Starting inertial load was determined as the load and movement velocity that maximized peak power output and was implemented at the beginning of week 3. Inertial load was again increased at the beginning of week 5 whereas movement velocity was increased at the beginning of weeks four and six. The systematic manipulation of inertial load and movement velocity ensured continued increases in power output throughout the exercise intervention. Participants were encouraged to perform the concentric action as fast as they could safely, then actively resisting the load during the eccentric phase before immediately initiating the next concentric action.

Following their flywheel resistance exercises, participants completed 30 minutes of aerobic exercise, or as much as they were able to tolerate, on a stationary cycle ergometer, at an intensity corresponding with 50-80% of their heart rate reserve.

### Health Education (EDU)

The participants assigned to the EDU group continued to receive standard care at the DC VAMC, in addition to weekly educational sessions for six weeks conducted by the research team on the importance of PA, exercise, and dietary habits. Participants were provided with the National Institute of Aging Exercise & Physical Activity handbook and asked to record weekly PA. Participants discussed their health goals with the research team, and the research team offered PA and exercise suggestions to help each participant achieve their goals.

### Statistical Analysis

Sample size was estimated to detect within- and between-group differences in knee extensor power output, assuming a 20% improvement following FRE+AE. For within-group differences using a paired t-test and 80% power for a two-tailed hypothesis at a 0.05 significance level, a total of 8 participants was estimated. For between-group differences using a two-sample independent t-test and 80% power for a two-tailed hypothesis at a 0.05 significance level, a total of 12 participants per group was estimated.

All data were normally distributed with equal variances according to the Kolmogorov-Smirnov test and the Levene’s test. For parametric data, an independent t-test were used to compare continuous variables and test the hypothesis that the FRE+AE group would exhibit greater improvements in neuromuscular and physical function outcomes than the EDU group. For nonparametric data, the Mann-Whitney test was used instead of the independent t-test. Paired t-test was used to test the hypothesis that FRE+AE would increase neuromuscular capacity and physical function in Veterans with eGFR 59-15 ml/min per 1.73m^2^. For nonparametric data, the Wilcoxon signed-rank test was used instead of the paired t-test. Exploratory analysis was performed by evaluating effect sizes (Hedge’s g) generated from changes in neuromuscular and physical function outcomes from baseline to six weeks between those meeting the weekly moderate intensity PA recommendations (Meets PA) to those below the moderate intensity PA recommendations (Low PA). Effect sizes were interpreted as small (<0.2), medium (0.5), and large (>0.8).

Pearson product-moment correlation coefficients (r) were used to assess bivariate associations between self-reported moderate intensity weekly PA and neuromuscular and physical function outcomes. Simple linear regression analyses were performed to test the hypothesis that weekly moderate intensity PA moderates neuromuscular and physical function adaptations following six weeks of FRE+AE in patients with CKD stages 3-4. In the model, post-intervention isokinetic power output at 180°·s^-1^, VL MT, and STS were regressed on baseline PA while adjusting for baseline isokinetic power output at 180°·s^-1^, VL MT, and STS. Baseline PA was mean-centered (i.e., each participant’s value minus the overall sample mean) prior to creating the interaction term. Centering was performed for two reasons: 1) to reduce multicollinearity between the main-effect terms and the interaction term (thereby stabilizing variance inflation factors and improving the interpretability of regression coefficients), and 2) to allow the main effect of group to represent the treatment effect at the average level of baseline PA rather than at an arbitrary zero value. Because the interaction effects were exploratory and intended to generate hypotheses for future research, we used an alpha level of p<0.10 for interaction terms.

## RESULTS

The average eGFR of the sample was 37.9 ml/min per 1.73 m^2^ with 20% classified as CKD stage 3a (eGFR 59-45 ml/min per 1.73 m^2^), 55% with CKD stage 3b (eGFR 44-30 ml/min per 1.73 m^2^), and 25% with CKD stage 4 (eGFR 29-15 ml/min per 1.73 m^2^). The average weekly self-reported PA of moderate intensity or greater was ∼509 minutes, and 55% of the total sample met the PA guideline of ≥150 minutes of moderate PA per week. According to BMI, 25% of the sample was classified as healthy weight (18.5-24.9), 40% as overweight (25-29.9), and 35% as obese (>30). No significant differences were observed in neuromuscular or physical function outcomes between those meeting the weekly guidelines for moderate-intensity PA and those below the recommendations (**Table 1**). Self-report PA level was not associated with any neuromuscular or physical function outcome (p>0.05).

**Table 1.**
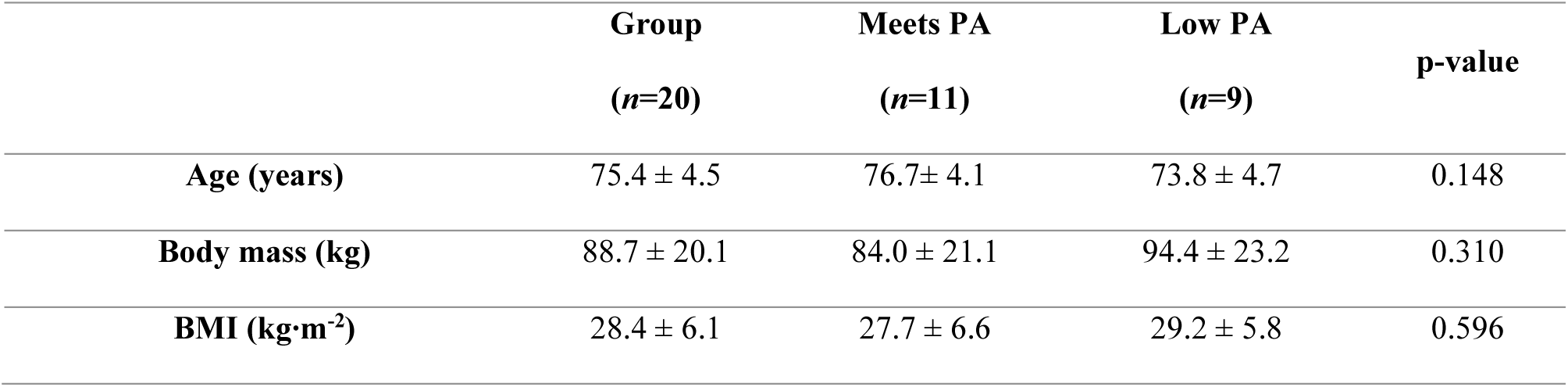
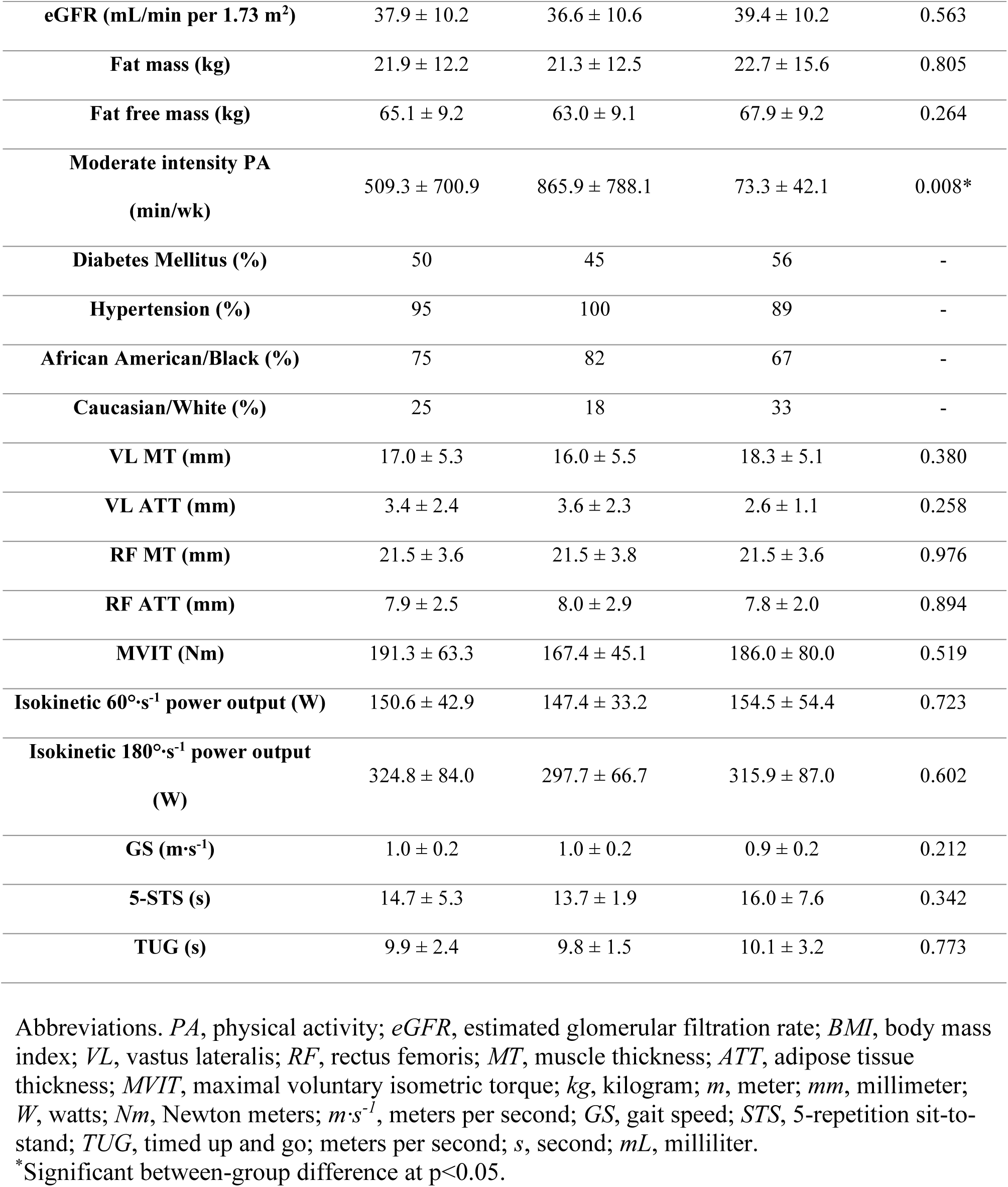
Participant characteristics according to physical activity status.

### FRE+AE Workload Progression

Inertial load increased significantly from week 3 to week 6 (0.031±0.011 kg·m^2^ vs 0.043±0.013 kg·m^2^, p<0.01) whereas contraction velocity did not change (12.3±2.1 rpm vs 12.0±1.5 rpm, p=0.415) (**Fig. 1**). Concentric power output and eccentric power output increased significantly from week 3 to week 6 (238.6±89.5 W vs 322.8±113.1 W, p<0.01; 221.0±81.1 W vs 300.0±96.1 W, p<0.01; respectively).

**Figure 1.**
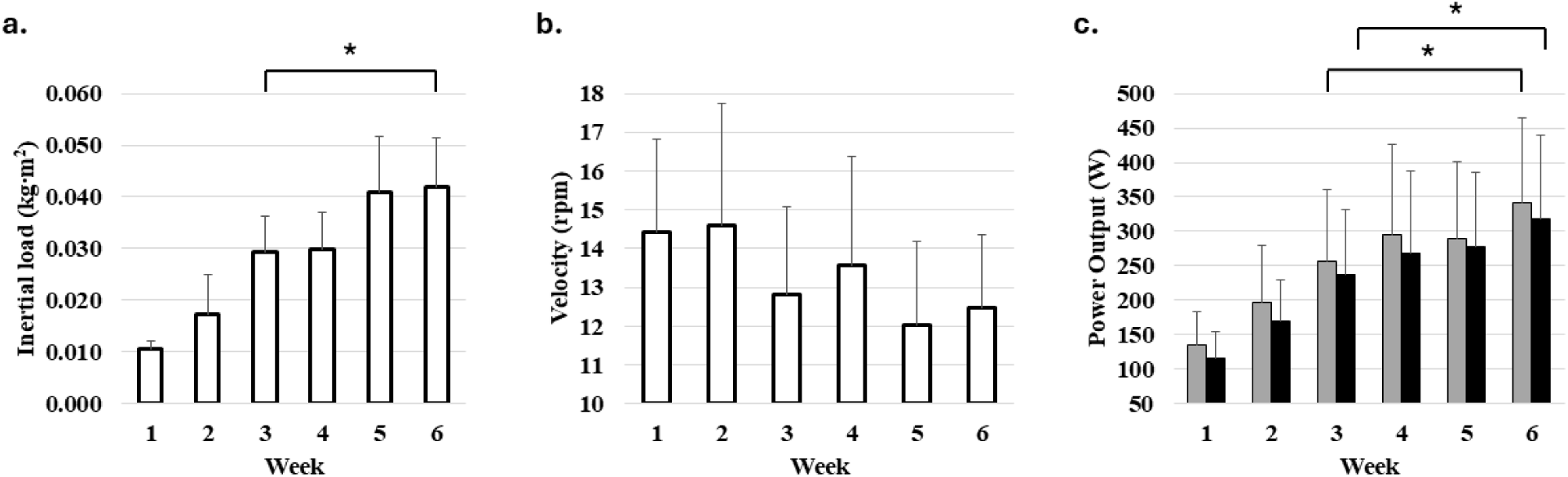
Depicts flywheel resistance exercise (FRE) progressions for the squat exercise over six weeks for (**a**) inertial load, (**b**) velocity, and (**c**) the resultant changes in concentric (gray bars) and eccentric (black bars) power output for the squat exercise. Data reflects group means and standard deviations. *Significantly difference between week 3 and 6 at p<0.01.

### Neuromuscular and Physical Function Adaptations

After six weeks of FRE+AE, significant increases in VL MT (p=0.030) and isokinetic power output 180°·s^-1^ (p=0.021) were observed (**Table 2**). For physical function, significant increases were seen for GS (p=0.001) and STS (p=0.012). In the EDU group, significant increases were noted in MVIT (p=0.045) and STS (p=0.046). Significant between-group differences were observed for change in VL MT (p=0.009) (**Fig. 2**) and gait speed (p=0.028) at six weeks (**Fig. 3**).

**Figure 2.**
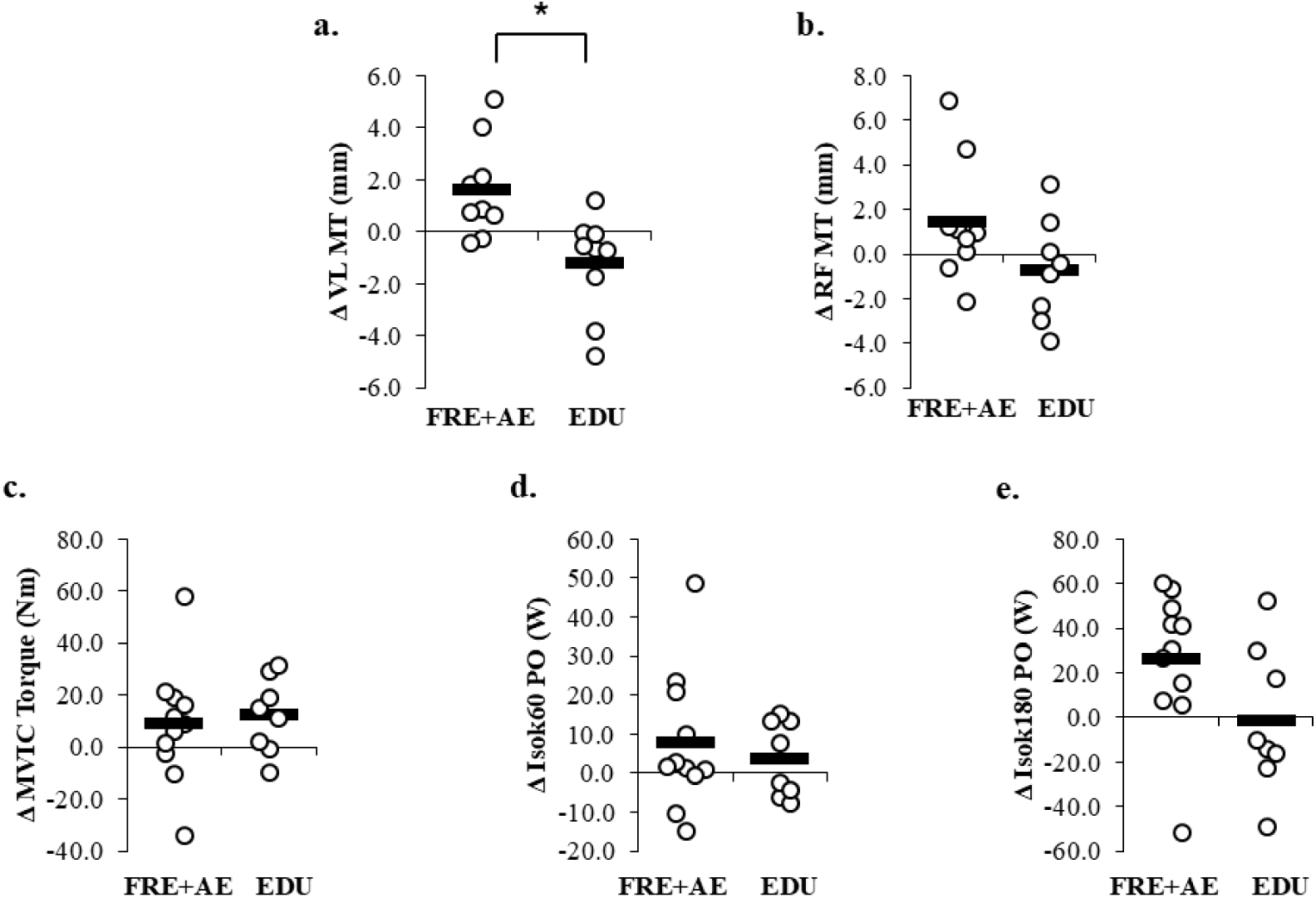
Depicts changes in (**a**) vastus lateralis muscle thickness (VL MT), (**b**) rectus femoris muscle thickness (RF MT), (**c**) knee extensor peak isometric torque (MVIT), (**d**) isokinetic power output at 60°·s^-1^, and (**e**)isokinetic power output at 180°·s^-1^ after six weeks of flywheel resistance exercise plus aerobic exercise (FRE+AE) or health education (EDU). Open circles represent individual participant responses and horizontal solid lines represent group means. *Significant between-group differences at p<0.05.

**Figure 3.**
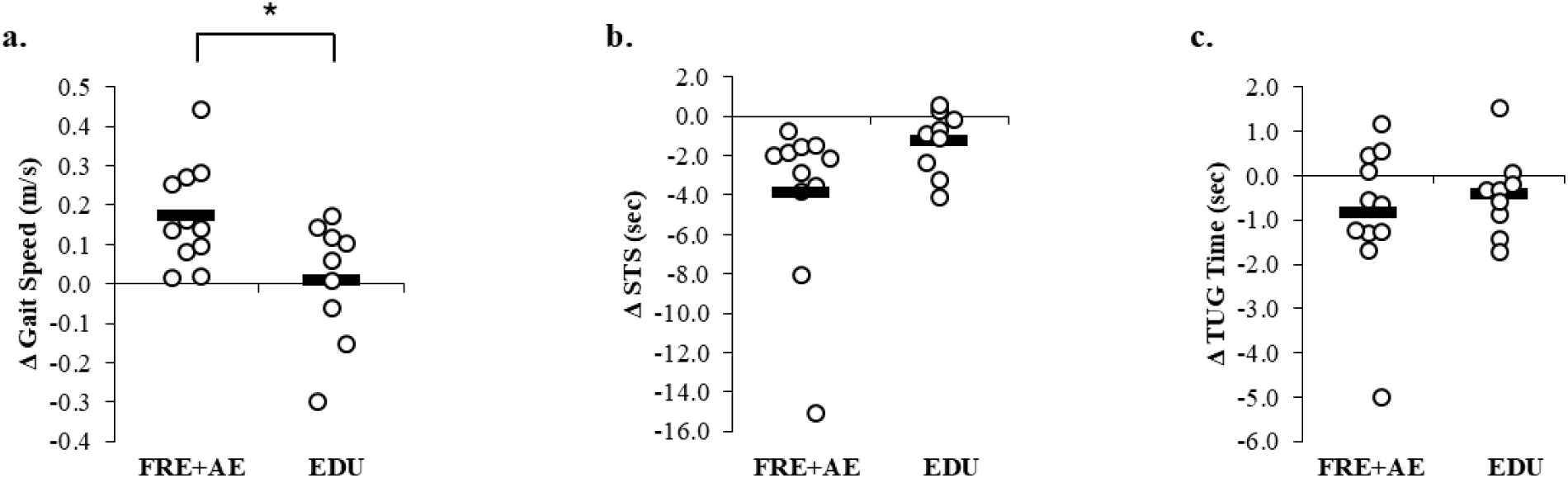
Depicts changes in (**a**) gait speed (GS), (**b**) time to complete five chair stands (STS), and (**c**) timed up and go (TUG) after six weeks of flywheel resistance exercise plus aerobic exercise (FRE+AE) or health education (EDU). Open circles represent individual participant responses and horizontal solid line represent group means. *Significant between-group differences at p<0.05.

**Table 2.** Changes in neuromuscular and physical function outcomes after six weeks of FRE+AE or EDU.

|  | FRE+AE (n=11) |  |  | EDU (n=9) |  |  |
| --- | --- | --- | --- | --- | --- | --- |
|  | Pre | Post | p-value | Pre | Post | p-value |
| VL MT (mm) | 16.5 ± 6.3 | 18.1 ± 6.0 | 0.030* | 17.6 ± 4.3 | 16.3 ± 3.7 | 0.094 |
| VL ATT (mm) | 3.2 ± 2.1 | 3.0 ± 2.1 | 0.524 | 3.1 ± 1.8 | 3.1 ± 1.9 | 0.761 |
| RF MT (mm) | 20.5 ± 3.3 | 22.0 ± 4.5 | 0.148 | 22.5 ± 3.8 | 21.8 ± 3.1 | 0.574 |
| RF ATT (mm) | 8.5 ± 3.0 | 9.2 ± 4.3 | 0.348 | 7.3 ± 1.6 | 7.0 ± 2.2 | 0.356 |
| MVIT (Nm) | 194.9 ± 61.4 | 203.9 ± 64.5 | 0.216 | 161.5 ± 54.3 | 173.9 ± 61.3 | 0.045* |
| Isokinetic Power Output<br>at 60°·s<sup>-1</sup> (W) | 156.2 ± 42.5 | 164.0 ± 37.0 | 0.174 | 150.8 ± 42.5 | 154.5 ± 40.8 | 0.313 |
| Isokinetic Power Output<br>at 180°·s<sup>-1</sup> (W) | 310.4 ± 66.3 | 336.6 ± 77.8 | 0.021* | 309.8 ± 89.2 | 308.6 ± 94.8 | 0.916 |
| GS (m·s<sup>-1</sup>) | 1.0 ± 0.2 | 1.1 ± 0.2 | 0.001* | 1.0 ± 0.2 | 1.0 ± 0.2 | 0.827 |
| STS (s) | 16.2 ± 6.7 | 12.3 ± 2.7 | 0.012* | 12.9 ± 2.1 | 11.6 ± 2.3 | 0.046* |
| TUG (s) | 9.9 ± 2.8 | 9.0 ± 2.1 | 0.123 | 10.0 ± 2.0 | 9.6 ± 1.4 | 0.224 |
| Moderate intensity PA<br>(min/wk) | 394.1 ± 589.6 | - | - | 650 ± 831.7 | - | - |

Abbreviations. *FRE+AE*, flywheel resistance exercise plus aerobic exercise; *EDU*, health education; *VL*, vastus lateralis; *RF*, rectus femoris; *MT*, muscle thickness; *ATT*, adipose tissue thickness; *MVIT*, maximal voluntary isometric contraction; *kg*, kilogram; *m*, meter; *mm*, millimeter; *W*, watts; *Nm*, Newton meters; *m·s<sup>-1</sup>*, meters per second; *GS*, gait speed; *STS*, 5-repetition sit-to-stand; *TUG*, timed up and go; meters per second; *s*, second; *mL*, milliliter.
\*Significant within-group difference at p<0.05.

### Bivariate Correlations Between Changes in Neuromuscular and Physical Function Outcomes

Change in VL MT was positively associated with change in isokinetic power output at 180°·s^-1^ (r=0.59, p=0.013) (**Fig. 4**). Change in STS and TUG were inversely associated with change in MVIT (r=-0.51, p=0.031 and r=-0.50, p=0.030) and change in isokinetic power output at 60°·s^-1^ (r=-0.64, p=0.003 and r=-0.50, p=0.031).

**Fig. 4.**
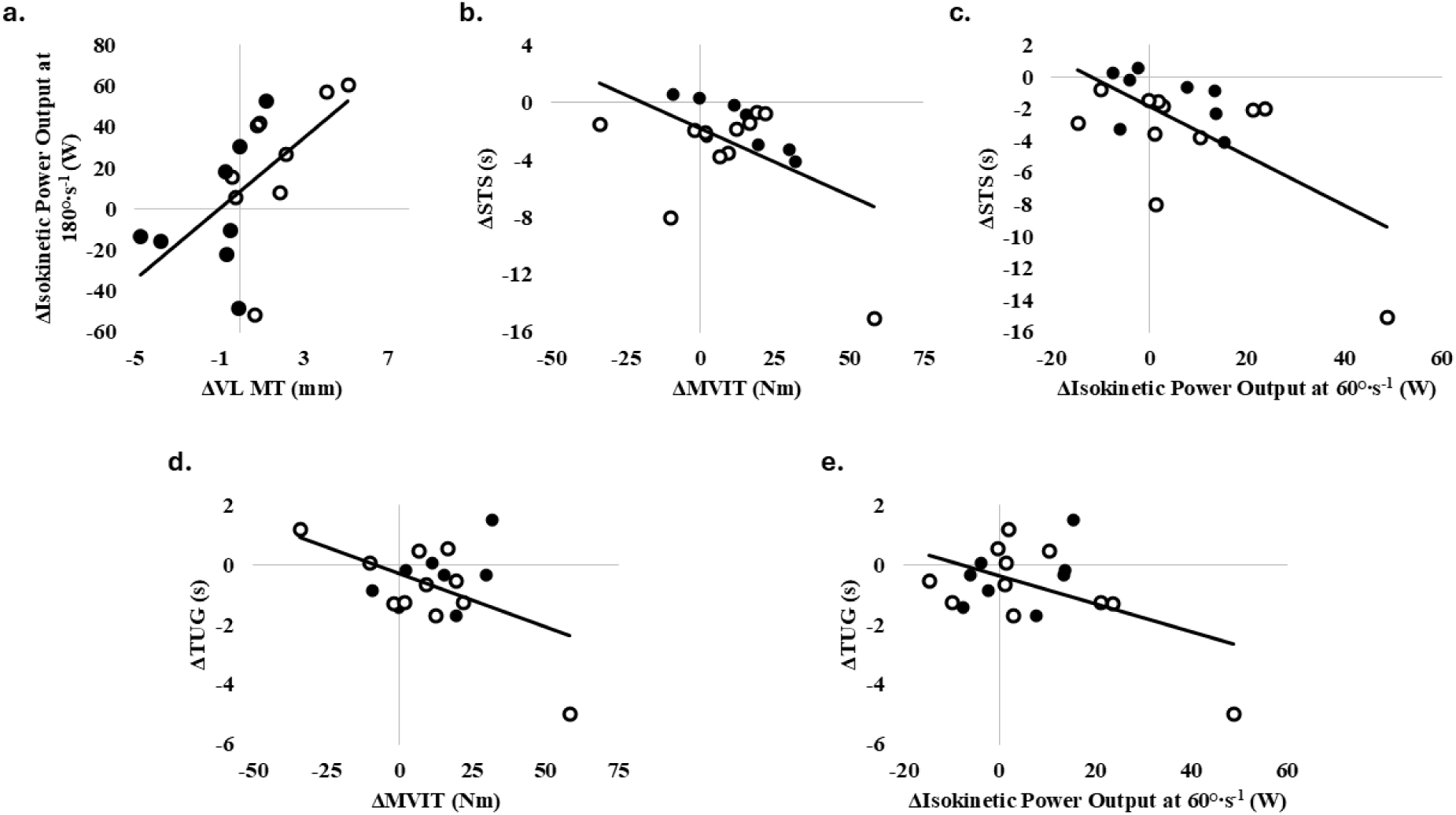
Bivariate correlations between (a) change in isokinetic power output at 180°·s^-1^ and change in vastus lateralis muscle thickness (ΔVL MT), (b) change in change in time to complete five sit-to-stands (ΔSTS) and change in maximal voluntary isometric torque (ΔMVIT), (c) ΔSTS and change in isokinetic power output at 60°·s^-1^, (d) change in timed up and go (ΔTUG) and ΔMVIT, and (e) ΔTUG and change in isokinetic power output at 60°·s^-1^. Open circles represent FRE+AE group and solid circles represent EDU group.

### Comparison of Changes in Neuromuscular Outcomes According to Physical Activity Level

In the FRE+AE group, increases were observed for all neuromuscular outcomes irrespective of PA classification (**Fig. 5**). The magnitude of change in VL MT was similar between PA classifications (Hedge’s g: Meets PA = 0.67, Low PA = 0.61; respectively). Greater magnitude of increase in RF MT (Hedge’s g: Meets PA = 0.26, Low PA = 0.52), isokinetic power output at 60°·s^-1^ (Hedge’s g: Meets PA = 0.25, Low PA = 0.44), and 180°·s^-1^ (Hedge’s g: Meets PA = 0.38, Low PA = 1.38) were observed in the Low PA classification whereas the magnitude of improvement for MVIT (Hedge’s g: Meets PA = 1.12, Low PA = 0.17) was greater in the Meets PA classification.

**Figure 5.**
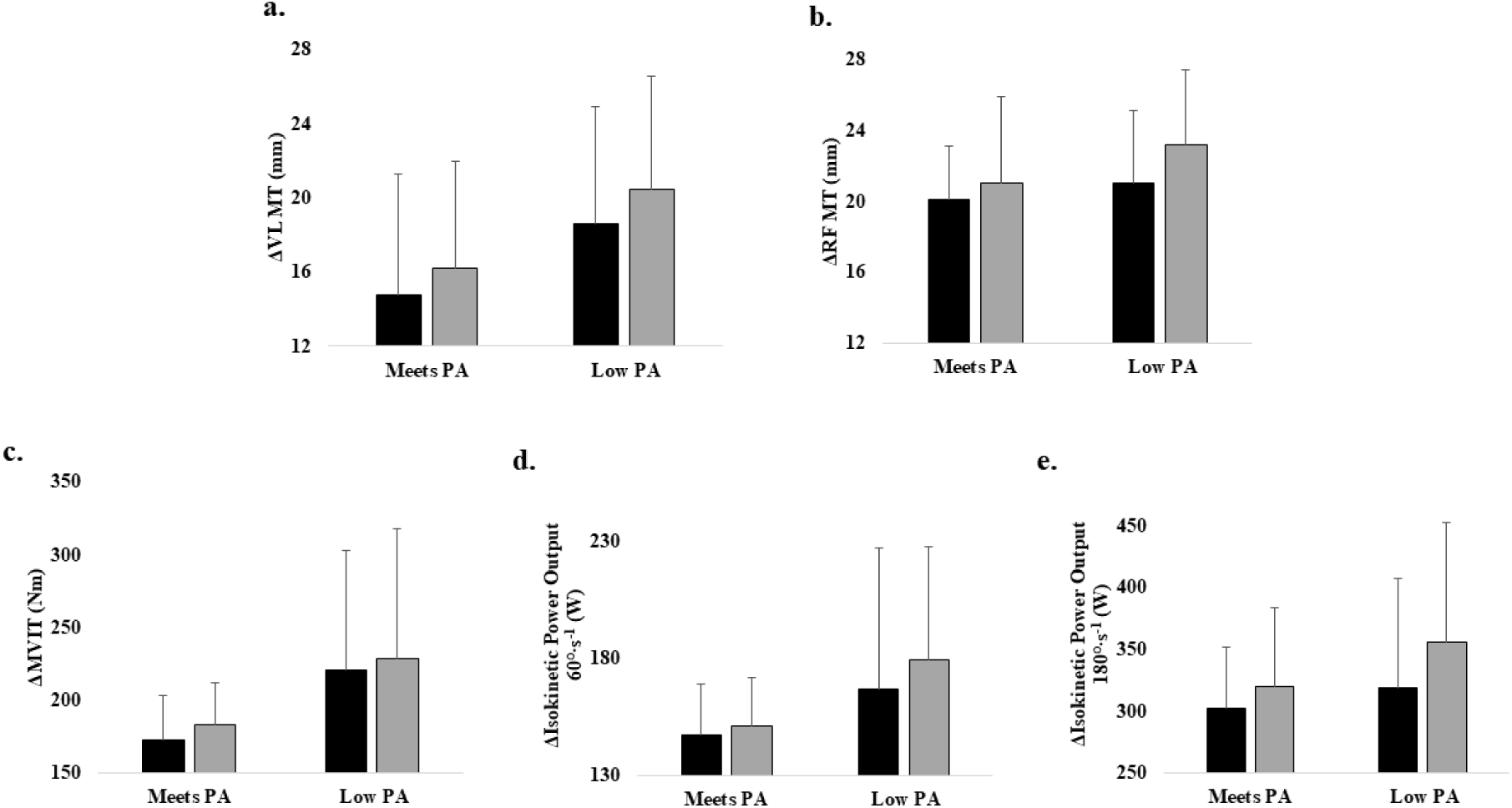
Comparison of changes in (**a**) vastus lateralis muscle thickness (VL MT), (**b**) rectus femoris MT (RF MT), (**c**) maximal voluntary isometric torque (MVIT), (**d**) isokinetic power output at 60°·s^-1^, and (**e**) isokinetic power output at 180°·/s^-1^ between participants meeting the moderate intensity physical activity recommendations (Meets PA, *n*=6) and participants below the moderate intensity physical activity recommendations (Low PA, *n*=5) after six weeks of flywheel resistance exercise plus aerobic exercise (FRE+AE). Black bars represent baseline and gray bars represent post-testing. Data are represented as means and standard deviation.

In the EDU group, decreases in VL MT (Hedge’s g: Meets PA = −0.40, Low PA = −0.55) and RF MT (Hedge’s g: Meets PA = −0.24, Low PA = −0.73) were observed irrespective of PA classification, with the magnitude of decline being greater in Low PA compared to Meets PA (**Fig. 6**). Those in the Meets PA classification also experienced greater magnitude of increase in MVIT (Hedge’s g: Meets PA = 1.05, Low PA = 0.34) and isokinetic power output at 60°·s^-1^ (Hedge’s g: Meets PA = 0.56, Low PA = 0.03) compared to the Low PA classification. Additionally, isokinetic power output at 180°·s^-1^ increased in the Meets PA classification but decreased in the Low PA classification (Hedge’s g: Meets PA = 0.36, Low PA = −0.46).

**Figure 6.**
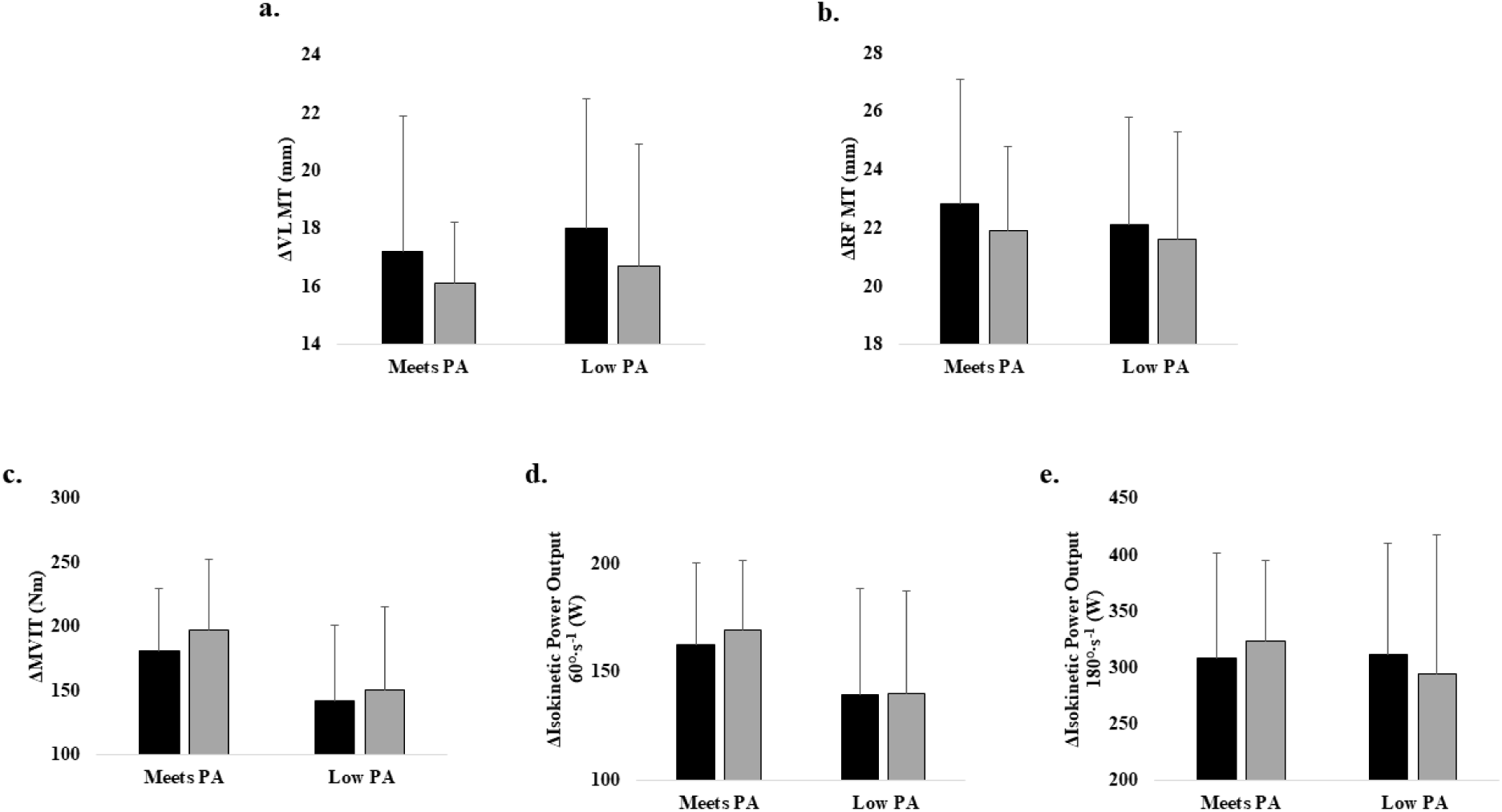
Comparison of changes in (**a**) vastus lateralis muscle thickness (VL MT), (**b**) rectus femoris MT (RF MT), (**c**) maximal voluntary isometric torque (MVIT), (**d**) isokinetic power output at 60°·s^-1^, and (**e**) isokinetic power output at 180°·/s^-1^ between participants meeting the moderate intensity physical activity recommendations (Meets PA, *n*=5) and participants below the moderate intensity physical activity recommendations (Low PA, *n*=4) after 6-weeks of health education (EDU). Black bars represent baseline and gray bars represent post-testing. Data are represented as means and standard deviation.

### Comparison of Changes in Physical Function Outcomes According to Physical Activity Level

In the FRE+AE group, increases were observed for all physical function outcomes irrespective of PA classification, with the magnitude of change being similar for both PA classifications (Hedge’s g: GS, Meets PA = 1.29, Low PA = 0.93; STS, Meets PA = −2.20, Low PA = −0.72; TUG, Meets PA = −0.47, Low PA = −0.42; respectively) (**Fig. 7**).

**Figure 7.**
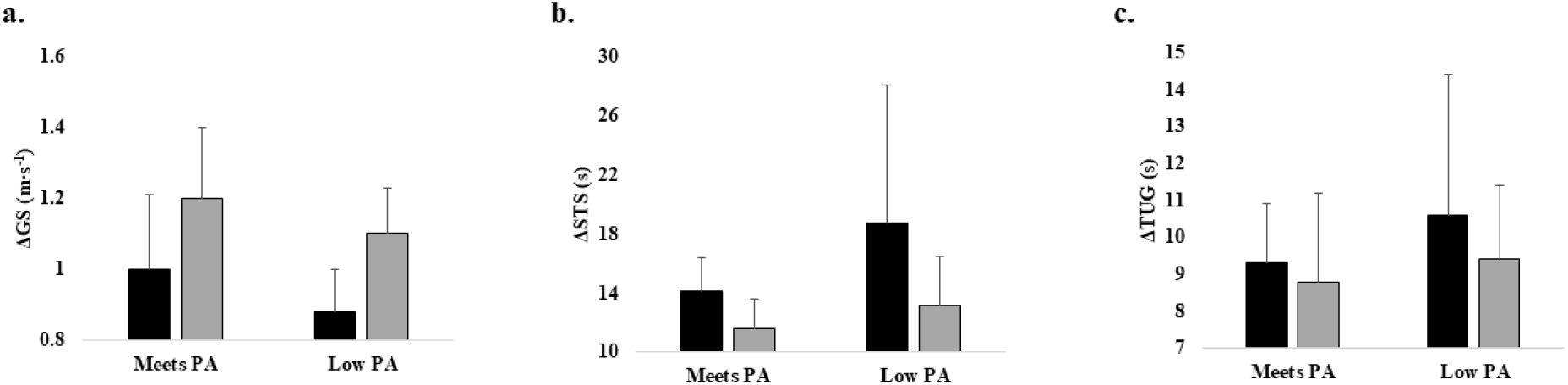
Comparison of changes in (**a**) gait speed (GS), (**b**) time to complete five sit-to-stands (STS), and (**c**) time to complete timed up-and-go (TUG) between participants meeting the moderate intensity physical activity recommendations (Meets PA, *n*=6) and participants below the moderate intensity physical activity recommendations (Low PA, *n*=5) after six weeks of flywheel resistance exercise plus aerobic exercise (FRE+AE). Black bars represent baseline and gray bars represent post-testing. Data are represented as means and standard deviation.

In the EDU group, the Low PA classification experienced null-to-small improvements in GS (Hedge’s g = 0.02), STS (Hedge’s g = −0.28), and TUG (Hedge’s g = −0.09) (**Fig. 8**). Conversely, large improvements were seen in STS (Hedge’s g: −1.16) and TUG (Hedge’s g = −0.78), and null improvements in GS (Hedge’s g = 0.02), for the Meets PA classification.

**Figure 8.**
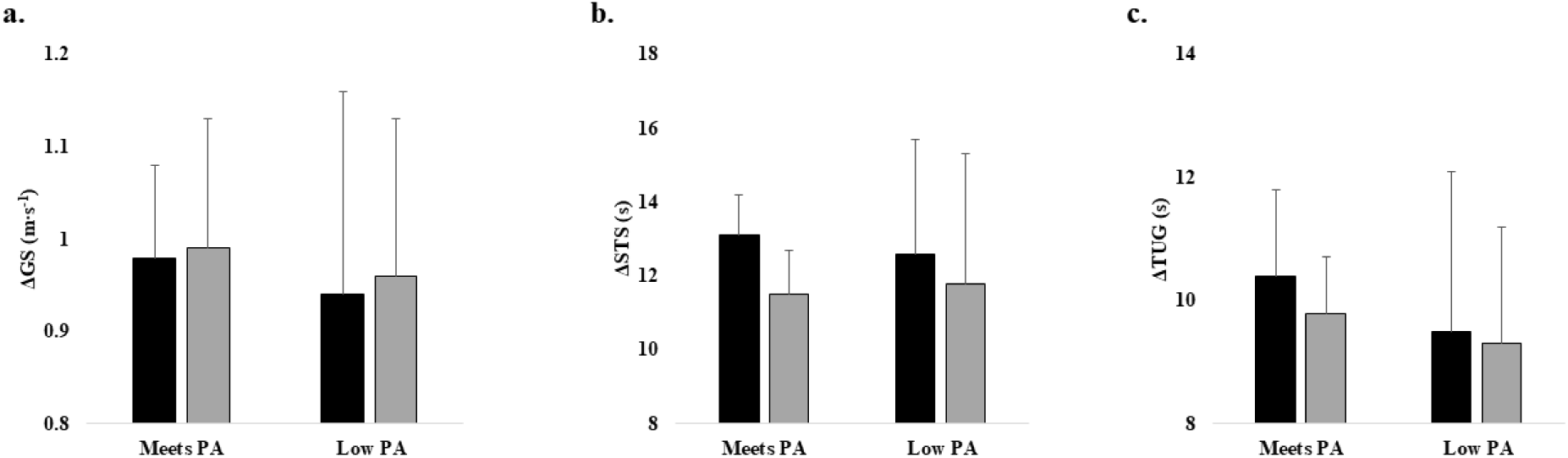
Comparison of changes in (**a**) gait speed (GS), (**b**) time to complete five sit-to-stands (STS), and (**c**) time to complete timed up-and-go (TUG) between participants meeting the moderate intensity physical activity recommendations (Meets PA, *n*=5) and participants below the moderate intensity physical activity recommendations (Low PA, *n*=4) after six weeks of health education (EDU). Black bars represent baseline and gray bars represent post-testing. Data are represented as means and standard deviation.

### Baseline Moderate Intensity PA as a Moderator of Adaptations

Findings from simple linear regression analyses revealed no statistical evidence that baseline PA was a moderator of the effects of FRE+AE on VL MT, isokinetic power output at 180°·s^-1^, or STS as indicated by the lack of model enhancement by the critical group × baseline PA interaction term (ΔR² ≤ 0.02, all p > 0.28) (**Table 3**).

**Table 3.**
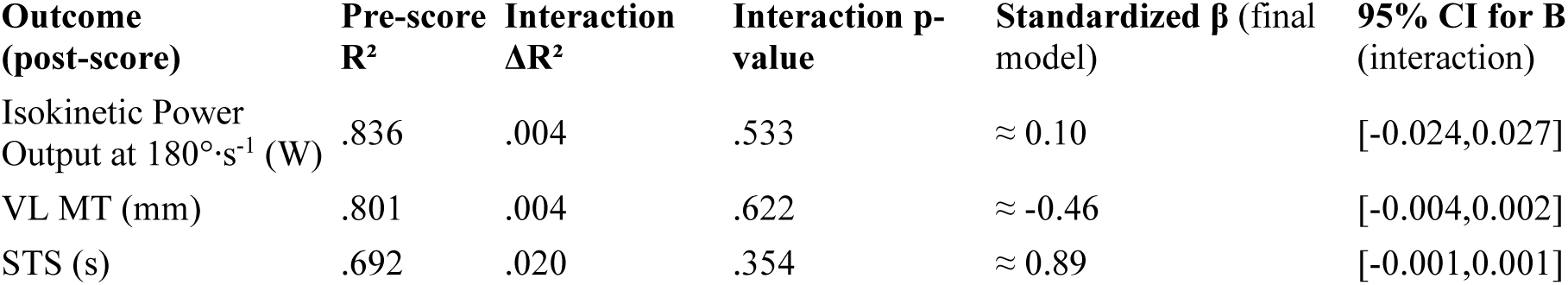
Simple linear regression model.

## DISCUSSION

Our findings demonstrate that six weeks of FRE+AE promote clinically meaningful improvements in VL MT, isokinetic power output at 180°·s^-1^, GS, and STS performance in older male Veterans with CKD stages 3-4. Exploratory analysis showed that Veterans not meeting the moderate intensity weekly PA recommendations experienced greater increases in RF MT and isokinetic power output (60°·s^-1^ and 180°·s^-^1) compared to physically active Veterans, with no differences in the magnitude of improvements in physical function between PA classifications. However, baseline self-reported PA level was not significantly associated with neuromuscular or physical function outcomes in older male Veterans with CKD stages 3-4, nor was it a moderator of neuromuscular or physical function adaptations to six weeks of FRE+AE. Overall, the results of the present study support FRE+AE as a feasible and effective strategy in older adults with CKD stages 3–4, irrespective of baseline habitual PA level.

The most common protocol for prescribing FRE consists of four sets of seven maximal-effort repetitions (Tesch *et al*., 2017). However, performing repetitions at maximal effort may not be safe or tolerable in older adults or certain clinical populations when engaging in an unaccustomed exercise modality. Our findings on FRE+AE build directly upon prior preliminary work demonstrating the feasibility and early benefits of eccentric-overload FRE in older Veterans with CKD using a progression scheme that manipulates inertial load and contraction velocity (Gollie *et al*., 2020). In that case series, short-term flywheel training improved torque capacity and physical function without adverse events, supporting the safety and exercise tolerance observed in the current study. The present RCT extends those results by incorporating a concurrent aerobic component, demonstrating additive neuromuscular and functional gains that are consistent with recent meta-analyses showing combined aerobic plus resistance exercise produces superior improvements in muscle strength, physical performance, and cardiorespiratory fitness compared with either modality alone in CKD populations (Liu *et al*., 2024). Notably, the magnitude of VL MT increase (approximately 10%) and isokinetic power gains (approximately 8% at 180°·s^-1^) mirror adaptations reported in healthy older adults following flywheel training (Sañudo *et al*., 2019; Hill *et al*., 2022) and suggest that the unique loading characteristics associated with rotational inertia are translatable to this clinical population. The enhanced adaptations in knee extensor power output at 180°·s^-1^, as compared to isometric torque and power output at 60°·s^-1^, highlight the benefits of progressing FRE contraction velocity, in addition to inertial load, on adaptations at higher contraction speeds. The ability to elicit adaptations in power output at higher contraction speeds may be particularly advantageous for enhancing physical function in selected clinical populations. Further research is required to determine the most optimal strategies for progressing inertial load and contraction velocity when targeting specific neuromuscular and function outcomes in diverse populations of healthy and unhealthy older adults.

The observed functional improvements underscore the clinical relevance of the FRE+AE intervention implemented in the present study. In the FRE+AE group, STS time improved from 16.2 s to 12.3 s, moving participants, on average, below the threshold associated with recurrent falls (>15 s) and sarcopenia-related lower-extremity function to levels consistent with average performance for men aged 70–79 years (Bohannon, 2006; Buatois *et al*., 2010; Cruz-Jentoft *et al*., 2019). The increase of 0.1 m/s in GS meets the threshold of substantial meaningful change in older adults (Perera *et al*., 2006). These gains occurred independent of baseline habitual PA classification and exceeded changes seen in the EDU group. Furthermore, increases in knee extensor MVIT and isokinetic power output (60°·s^-^1) were associated with reductions in time to complete five STS and TUG. These data align with broader evidence that combined resistance and aerobic training improves objective physical function in patients with non-dialysis CKD (Hellberg *et al*., 2019; Villanego *et al*., 2020; Weiner *et al*., 2023; Correa *et al*., 2025).

Robust evidence supports the importance of PA for maintaining health across the lifespan (Myers *et al*., 2015; Booth *et al*., 2017; Lobelo *et al*., 2018; Kraus *et al*., 2019; Ramakrishnan *et al*., 2021). However, studies examining the association between PA and neuromuscular outcomes have been inconclusive. Several cross-sectional studies found a lack of association between PA level and muscle characteristics (Morie *et al*., 2010; Leblanc *et al*., 2015; Perkin *et al*., 2018) whereas others report a positive association between PA and muscle size and strength (Rantanen *et al*., 1998; Hunter *et al*., 2000; Ikezoe *et al*., 2011; Ramsey *et al*., 2021). The same is true for longitudinal studies, with some investigations finding an association between PA level and changes in strength and power (Rantanen & Heikkinen, 1998; Alcazar *et al*., 2023) while others did not (Hughes *et al*., 2001, 2002; Buchman *et al*., 2007). Possible explanations for the discrepancies across studies include differences in sex, age, racial and ethnic distribution across studies, methods used to capture PA levels and neuromuscular outcomes, skeletal muscle(s) assessed, types and intensity of activities performed, and homogeneity of sample populations.

Our results suggest that neuromuscular and functional adaptations are achieved using FRE+AE in older men with non-dialysis CKD regardless of baseline habitual PA status. We observed comparable adaptations between PA classifications for VL MT following six weeks of FRE+AE, whereas the magnitudes of adaptations with exercise were greater for RF MT and isokinetic power output (60°·s^-1^ and 180°·s^-^1) output in the Low PA versus Meets PA classifications. These findings seem to, in part, align with the concept of diminishing returns and are in partial support of our hypothesis that greater adaptations would be observed in participants with reduced PA levels. Conversely, in the EDU group, those meeting the PA recommendations for moderate-intensity PA experience greater increases in isometric torque and power output at 60°·s-1, while attenuating declines in VL MT, RF MT, and power output at 180°·s^-1^ compared to those not meeting the PA recommendations. Thus, in our study population, being physically active did not exert an influence on neuromuscular adaptations achieved with FRE+AE but may attenuate declines in neuromuscular characteristics in the absence of structured exercise. This latter point is critical, as declines in PA and physical function are known to occur in patients with advanced CKD, and are further exacerbated with the transition to dialysis (Rampersad *et al*., 2021).

The magnitudes of improvements for physical function in the FRE+AE group was comparable between PA classifications, demonstrating large-to-moderate effects across all functional outcomes. In the EDU group, however, the improvements in STS and TUG were greater in the Meets PA classification compared to the Low PA classification, with no differences between PA classifications for GS. The trends observed for adaptations in physical function according to PA level in the present study are consistent with those in the existing literature (Buchman *et al*., 2007; Morey *et al*., 2008; Landi *et al*., 2018; Edholm *et al*., 2019; Ramsey *et al*., 2021; Rampersad *et al*., 2021; Cefis *et al*., 2025). The greater influence of PA on physical function, as compared to muscle size and strength, may be reflective of the specificity of activities performed. For example, walking is the most common form of PA performed by adults 50 years and older (Kuzmik *et al*., 2023). These findings have clinical implications for older adults with

CKD. While the KDIGO 2024 guidelines strongly recommend moderate-intensity PA (≥150 min/week), guidance on exercise modality and resistance training remains limited. Our results, together with recent evidence showing superior benefits of combined aerobic and resistance exercise in non-dialysis CKD (Liu *et al*., 2024), support incorporating structured combined programs into routine care. Interventions such as flywheel resistance exercise plus aerobic exercise provide a feasible, supervised strategy to achieve meaningful neuromuscular and functional gains regardless of baseline PA level, helping address the high burden of physical dysfunction and sarcopenia in this population.

Limitations of our study include the small sample size, exclusive enrollment of male Veterans (which limits generalizability to women and non-Veteran populations), the magnitude and six week duration of the intervention, and reliance on self-reported aerobic PA. Although the 7-day PAR provided a practical and standardized assessment of habitual activity, self-reported measures are inherently susceptible to recall bias, particularly among older adults. Participants often overestimate moderate-intensity activity and have difficulty accurately recalling the duration and intensity, which may have reduced our ability to detect subtle moderation effects or led to misclassification of individuals near the 150 min/week threshold. While no serious adverse events were reported, longer-term follow-up is needed to determine sustainability of the exercise adaptations and potential effects on clinical outcomes such as falls, hospitalization, or CKD progression. In addition, the homogeneous and relatively preserved baseline neuromuscular profile may also have masked potential moderation effects of PA on changes in muscle thickness, strength, and physical function.

## CONCLUSIONS

In conclusion, six weeks of FRE+AE elicited clinically meaningful neuromuscular and physical function improvements in older male Veterans with CKD stages 3–4; irrespective of habitual PA level. Data from the EDU group further underscores the benefits of being physically active for attenuating declines in neuromuscular health and physical function absent structured exercise.

Collectively, these results support the incorporation of FRE+AE as an alternative approach to clinical exercise prescriptions for the CKD population and highlight the need for longer-duration trials that incorporate objective PA monitoring, diverse, more representative cohorts, mechanistic outcomes, and larger sample sizes to optimize exercise programming and confirm long-term health benefits and safety.

## Data Availability

All data produced in the present study are available upon reasonable request to the authors

## Acknowledgements

This study was funded by the United States Department of Veterans Affairs, Office of Research and Development, Rehabilitation Research, Development, and Translation (RRDT) Section Career Development Award (CDA-2) 1IKRX003423 to J.M.G. and RRDT Senior Research Career Scientist Award IK6 RX003977 to A.S.R. This material is the result of work supported with resources and the use of facilities at the Washington DC VA Medical Center. The results of the study are presented clearly, honestly, and without fabrication, falsification, or inappropriate data manipulation. Results of the present study do not constitute endorsement by the United States Department of Veterans Affairs.

## Notes

### Competing Interest Statement

The authors have declared no competing interest.

### Clinical Trial

NCT04397159

### Author Declarations

Institutional Review Board and Research & Development Committee of the Washington DC VA Medical Center gave ethical approval of this work.

